# Transmission of SARS-CoV-2 Omicron VOC subvariants BA.1 and BA.2: Evidence from Danish Households

**DOI:** 10.1101/2022.01.28.22270044

**Authors:** Frederik Plesner Lyngse, Carsten Thure Kirkeby, Matthew Denwood, Lasse Engbo Christiansen, Kåre Mølbak, Camilla Holten Møller, Robert Leo Skov, Tyra Grove Krause, Morten Rasmussen, Raphael Niklaus Sieber, Thor Bech Johannesen, Troels Lillebaek, Jannik Fonager, Anders Fomsgaard, Frederik Trier Møller, Marc Stegger, Maria Overvad, Katja Spiess, Laust Hvas Mortensen

## Abstract

The Omicron SARS-CoV-2 variant of concern (VOC lineage B.1.1.529), which became dominant in many countries during early 2022, includes several subvariants with strikingly different genetic characteristics. Several countries, including Denmark, have observed the two Omicron subvariants: BA.1 and BA.2. In Denmark the latter has rapidly replaced the former as the dominant subvariant.

Based on nationwide Danish data, we estimate the transmission dynamics of BA.1 and BA.2 following the spread of Omicron VOC within Danish households in late December 2021 and early January 2022.

Among 8,541 primary household cases, of which 2,122 were BA.2, we identified a total of 5,702 secondary infections among 17,945 potential secondary cases during a 1-7 day follow-up period. The secondary attack rate (SAR) was estimated as 29% and 39% in households infected with Omicron BA.1 and BA.2, respectively.

We found BA.2 to be associated with an increased susceptibility of infection for unvaccinated individuals (Odds Ratio (OR) 2.19; 95%-CI 1.58-3.04), fully vaccinated individuals (OR 2.45; 95%-CI 1.77-3.40) and booster-vaccinated individuals (OR 2.99; 95%-CI 2.11-4.24), compared to BA.1. We also found an increased transmissibility from unvaccinated primary cases in BA.2 households when compared to BA.1 households, with an OR of 2.62 (95%-CI 1.96-3.52). The pattern of increased transmissibility in BA.2 households was not observed for fully vaccinated and booster-vaccinated primary cases, where the OR of transmission was below 1 for BA.2 compared to BA.1.

We conclude that Omicron BA.2 is inherently substantially more transmissible than BA.1, and that it also possesses immune-evasive properties that further reduce the protective effect of vaccination against infection, but do not increase its transmissibility from vaccinated individuals with breakthrough infections.

## 2 Introduction

The current pandemic with SARS-CoV-2 is characterized by continuous emergence of new variants taking over from previous variants as a result of natural selection (Darwin, 1859). Most recently, the Omicron variant of concern (VOC), Pango lineage B.1.1.529, has become the most prevalent in most countries in Europe as well as the rest of the world (Ritchie & Roser, 2020). Of the previously identified Omicron subvariants (Mullen et al., 2022; SSI, 2022), three subvariants have been detected in Denmark, namely BA.1.1, BA.1 and BA.2, where the latter two by far have been the most abundant. BA.1 and BA.2 differ by approximately 40 mutations (Majumdar & Sarkar, 2021) in addition to a key deletion at position 69-70 in the spike region of BA.1 compared to BA.2 (Public Health England, 2022; Chowdhury et al., 2022).

By 1 January 2022, BA.2 accounted for 5% of all subvariants found in England, with an ongoing increase in this proportion (Public Health England, 2022). BA.1 was first detected in Denmark on 25 November 2021, and BA.2 was first detected on 5 December 2021. Since then, the prevalence of BA.2 has been increasing faster than that of BA.1. In week 52 of 2021, BA.2 accounted for around 20% of all Danish SARS-CoV-2 cases; in week 2 in 2022 this had increased to around 45%, indicating that BA.2 carries an advantage over BA.1 within the highly vaccinated population of Denmark. The RT-PCR test used in Denmark does not target the S-gene deletion to detect Omicron cases, but instead targets the spike position L452 Wt (Spiess et al., 2021). Thus in the current set-up, Danish RT-PCR data cannot distinguish between BA.1 and BA.2. However, whole genome sequencing (WGS) is conducted routinely in Denmark (www.covid19genomics.dk), providing the opportunity to identify and differentiate between BA.1 and BA.2.

We have previously used a model of household transmission to quantify the transmissibility of VOCs, and applied this model to show that the Omicron VOC had an advantage over the Delta VOC due to immune evasiveness (Lyngse et al., 2021b).

The increasing numbers of BA.2 cases justify the questions we address in this study; 1) Is there a difference in the household transmission patterns between Omicron VOC subvariant BA.1 and BA.2; and 2) if there is a difference, is it due to a difference in susceptibility, transmissibility, or both, and could this indicate a difference in immune evasiveness between the subvariants?

## 3 Methods

### 3.1 Study design and participants

In this study, we used Danish register data comprising all individuals in Denmark. We linked all individuals to households by their personal identification number. We only included households with 2-6 members to exclude care facilities etc. We linked this with information on all antigen and RT-PCR tests for SARS-CoV-2 from the Danish Microbiology Database (MiBa; Schønning et al. (2021)), and records in the Danish Vaccination Register (Krause et al., 2012). We used data on primary cases from 20 December 2021 to 11 January 2022, and allowed a 7-day follow-up period for potential secondary cases, i.e. until 18 January 2022.

A primary case was defined as the first individual in a household testing positive with an RT-PCR test within the study period and being identified with the Omicron VOC BA.1 or BA.2 by WGS. We followed all tests of other household members in the follow-up period. A positive secondary case was defined by either a positive RT-PCR test or a positive antigen test (Jakobsen et al., 2021). Households were categorized as BA.1 or BA.2 households depending on the WGS result of the sample from the primary case.

In the study period, a total of approximately 25,000 mid- and high-quality SARS-CoV-2 genomes were produced (appendix Table 4) at the time of analysis. Briefly, sequencing of positive SARS-CoV-2 samples was performed using short read Illumina technology (Illumina) with the Illumina COVIDSeq Test kit (Illumina). The library preparation was performed as described by the manufacturer with spike-in of amplicon 64, 70 and 74 from the ARTIC v3 amplicon sequencing panel (https://artic.network). Samples were sequenced on either the NextSeq or NovaSeq platforms (Illumina). Consensus sequences were called using an in-house implementation of IVAR (version 1.3.1) with a custom BCFtools (Li, 2011) command for consensus calling. The resulting consensus sequences were considered for variant calling when containing <3,000 ambiguous sites including N’s. Variants were called using Pangolin (version 3.1.17) with PangoLEARN assignment algorithm (version 2022-01-05) on the consensus sequences (O’Toole et al., 2021).

The vaccination status of all individuals was classified into three groups following Lyngse et al. (2021b): i) unvaccinated (including partially vaccinated individuals); ii) fully vaccinated (defined by the vaccine used, Comirnaty (Pfizer/BioNTech): 7 days after second dose; Vaxzevria (AstraZeneca): 15 days after second dose; Spikevax (Moderna): 14 days after second dose; Janssen (Johnson & Johnson): 14 days after vaccination, and 14 days after the second dose for cross vaccinated individuals) or 14 days after previous infection; or iii) booster-vaccinated, defined by 7 days after the booster vaccination, (Pfizer, 2021; Bomze et al., 2021). By 22 December 2021, of all vaccinated individuals in Denmark, 85% were vaccinated with Comirnaty, 14% with Spikevax, 1% with Janssen, and approximately 0% with AstraZeneca (SSI, 2021).

### 3.2 Statistical analyses

The secondary attack rate (SAR) was defined as the proportion of potential secondary cases within the same household that tested positive between 1-7 days following the positive test of the primary case in that household. We estimated the adjusted odds ratios (OR) for infection in a multivariable logistic regression model. The outcome variable in this model was the binary test result of each potential secondary case. We used the sub-variant as an explanatory variable as well as fixed effects for other potentially confounding variables; age and sex of the primary case, age and sex of the potential secondary case, household size (2-6 members), and primary case sample date to control for time related effects. To test if the subvariants behaved differently depending on the immune status of the primary cases (i.e. different transmissibility) and the potential secondary cases (i.e. different susceptibility), we included interactions between household subvariant and vaccination status of the primary cases and the potential secondary cases, respectively.

To investigate the robustness of the results, we conducted a number of additional sensitivity analyses, which can be found in the appendix. Here, we also describe the distribution of BA.1 and BA.2 cases over the study period and the characteristics of samples selected for WGS. We also provide statistics for a 14-day follow-up period. Additionally, we provide measures of model fit and estimates under a number of alternative specifications of the logistic regression model to assess the robustness of the findings, as well as a more detailed investigation of the pairwise OR between vaccination groups for each subvariant.

### 3.3 Ethical statement

This study was conducted using data from national registers only. According to Danish law, ethics approval is not needed for this type of research. All data management and analyses were carried out on the Danish Health Data Authority’s restricted research servers with project number FSEID-00004942. The study only contains aggregated results and no personal data. The study is, therefore, not covered by the European General Data Protection Regulation (GDPR).

### 3.4 Data availability

The data used in this study are available under restricted access due to Danish data protection legislation. The data are available for research upon reasonable request to The Danish Health Data Authority and Statens Serum Institut and within the framework of the Danish data protection legislation and any required permission from Authorities. We performed no data collection or sequencing specifically for this study. Consensus genome data from the Danish cases are routinely shared publicly at GISAID (www.gisaid.org).

## 4 Results

We identified 2,122 households with BA.2 comprising a total of 4,587 potential secondary cases, of which 1,792 tested positive within 7 days, resulting in a SAR of 39%. Similarly, we identified 6,419 households with BA.1 comprising a total of 13,358 potential secondary cases, of which 3,910 tested positive, resulting in a SAR of 29%. The distributions of age, sex, household size and vaccination status in primary and potential secondary cases were broadly comparable between BA.1 and BA.2 households (Table 1).

**Table 1:** Summary statistics

|  | Omicron - BA.2 |  |  |  | Omicron - BA.1 |  |  |  |
| --- | --- | --- | --- | --- | --- | --- | --- | --- |
|  | Primary Cases | Potential Secondary Cases | Positive Secondary Cases | SAR (%) | Primary Cases | Potential Secondary Cases | Positive Secondary Cases | SAR (%) |
| **Total, N** | 2,122 | 4,587 | 1,792 | 39 | 6,419 | 13,358 | 3,910 | 29 |
| **Sex, %** |  |  |  |  |  |  |  |  |
| Male | 49 | 50 | 49 | 38 | 49 | 50 | 48 | 28 |
| Female | 51 | 50 | 51 | 40 | 51 | 50 | 52 | 31 |
| **Age, %** |  |  |  |  |  |  |  |  |
| 0-10 | 8 | 17 | 19 | 42 | 5 | 17 | 19 | 33 |
| 10-20 | 25 | 18 | 16 | 34 | 23 | 19 | 17 | 26 |
| 20-30 | 23 | 14 | 13 | 38 | 27 | 15 | 16 | 31 |
| 30-40 | 15 | 12 | 18 | 56 | 15 | 11 | 15 | 42 |
| 40-50 | 11 | 18 | 17 | 38 | 12 | 18 | 17 | 27 |
| 50-60 | 11 | 14 | 12 | 32 | 11 | 14 | 10 | 22 |
| 60-70 | 4 | 4 | 3 | 35 | 5 | 4 | 4 | 25 |
| 70+ | 3 | 2 | 2 | 30 | 3 | 2 | 2 | 26 |
| **Household size, %** |  |  |  |  |  |  |  |  |
| 2 | 37 | 17 | 19 | 44 | 40 | 19 | 22 | 33 |
| 3 | 24 | 23 | 21 | 37 | 25 | 24 | 22 | 27 |
| 4 | 26 | 36 | 38 | 41 | 23 | 34 | 35 | 31 |
| 5 | 10 | 18 | 16 | 34 | 9 | 18 | 17 | 28 |
| 6 | 2 | 6 | 5 | 35 | 2 | 5 | 4 | 23 |
| **Immunity, %** |  |  |  |  |  |  |  |  |
| Unvaccinated | 21 | 26 | 29 | 43 | 16 | 26 | 30 | 34 |
| Fully vaccinated / previous infection | 52 | 42 | 44 | 41 | 59 | 44 | 47 | 31 |
| Booster vaccinated | 26 | 32 | 27 | 34 | 25 | 30 | 23 | 22 |

Notes: The secondary attack rate (SAR) is expressed as a percentage (%). Total numbers (N) is presented in the first row. Summary statistics based on primary cases are shown separately from summary statistics of potential secondary cases, positive secondary cases and SAR. The raw numbers (N) for each category is presented in appendix Table 5. The summary statistics stratified by the primary case level are presented in appendix Tables 6 and 7.

Within 7 days of the identification of the primary cases, 84% of the potential secondary cases had been tested once, regardless of subvariant, and 60-61% had been tested twice (Figure 1, panel a). In households infected with the Omicron BA.2 (red), the SAR was 8% on day 1 and 39% on day 7. Similarly, in households infected with BA.1 (blue), the SAR was 6% and 29%, respectively.

**Figure 1:**
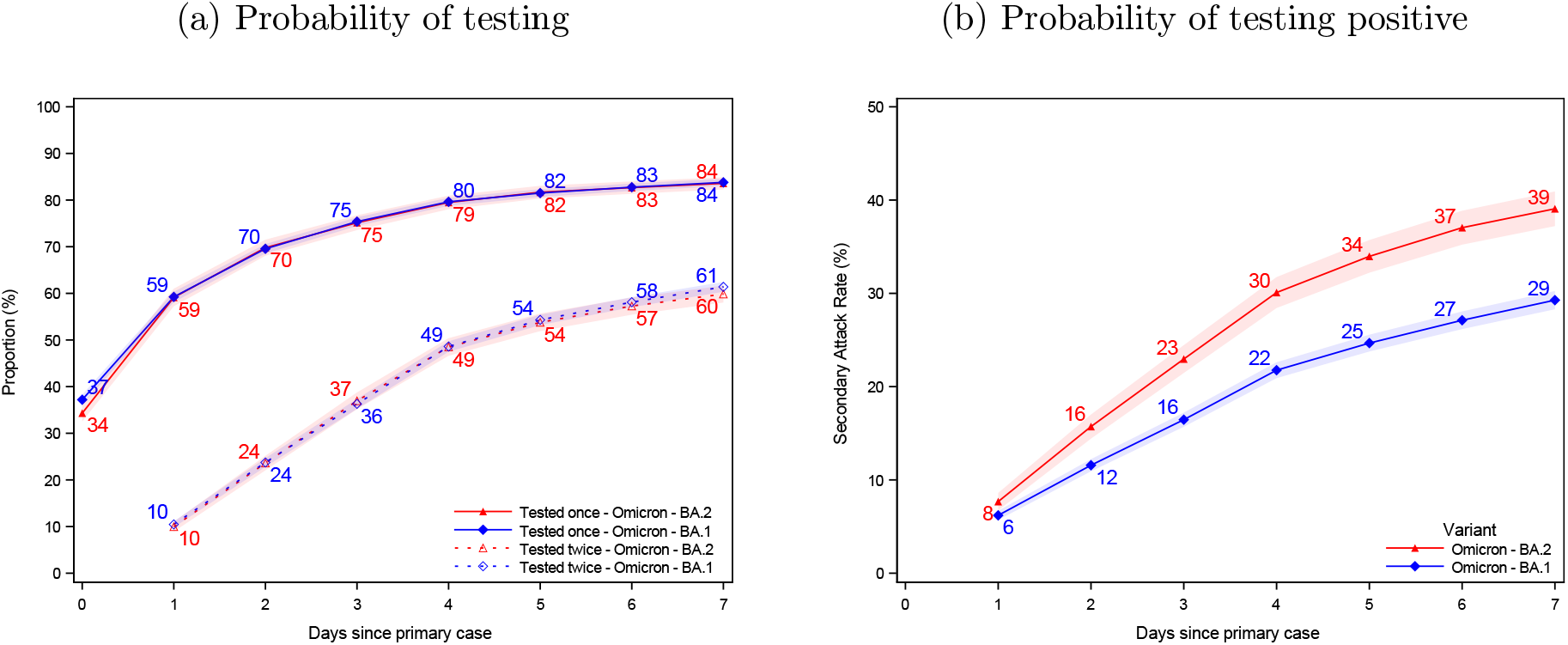
Probability of being tested and testing positive Notes: Panel (a) shows the probability of potential secondary cases being tested after a primary case has been identified within the household. Panel (b) shows the probability of potential secondary cases that test positive subsequently to a primary case being identified within the household. Note that the latter is not conditional on being tested, i.e. the denominator contains test-negative individuals and untested individuals. The x-axes show the days since the primary case tested positive, and the y-axes show the proportion of individuals either being tested (a) or testing positive (b) with either antigen or RT-PCR tests, stratified for the subvariant of the primary case. The SAR for each day according to the subvariant primary case can be read directly from panel (b). For example, the SAR on day 7 is 39% for BA.2 (red) and 29% for BA.1 (blue), whereas the SAR on day 4 is 30% and 22%, respectively. The shaded areas show the 95% confidence bands clustered on the household level. Appendix Figure 3 presents the two panels with a 14 day follow-up period. Appendix Figure 4 presents the 14 day SAR for Omicron BA.1, BA.2, and Delta VOC, as well as those without a known variant.

We observed a general gradient in both Omicron BA.1 and BA.2 households such that the susceptibility of potential secondary cases was highest among the unvaccinated and lowest among the booster vaccinated, but the effect of vaccination appeared to be lower for Omicron BA.2 than for BA.1 (see Table 2, and the interactions in Figure 6). We observed lower transmissibility in both BA.1 and BA.2 households when the primary case was booster vaccinated rather than fully vaccinated.

**Table 2:** Effect of Vaccination

|  | Susceptibility |  | Transmissibility |  |
| --- | --- | --- | --- | --- |
|  | Omicron BA.2 households | Omicron BA.1 households | Omicron BA.2 households | Omicron BA.1 households |
| Unvaccinated | 1.10<br>(0.92-1.32) | 1.23<br>(1.09-1.40) | 1.21<br>(0.97-1.50) | 0.93<br>(0.80-1.08) |
| Fully vaccinated | ref<br>(.) | ref<br>(.) | ref<br>(.) | ref<br>(.) |
| Booster vaccinated | 0.80<br>(0.67-0.94) | 0.65<br>(0.58-0.73) | 0.79<br>(0.64-0.98) | 0.77<br>(0.70-0.88) |
| Number of observations | 17,945 | 17,945 | 17,945 | 17,945 |
| Number of households | 8,541 | 8,541 | 8,541 | 8,541 |

Relative to Omicron BA.1 households, we found an increased susceptibility for both unvaccinated (OR 2.19; 95%-CI 1.58-3.04), fully vaccinated (OR 2.45; 95%-CI 1,77-3,40) and booster-vaccinated individuals (OR 2.99; 95%-CI 2.11-4.24) in BA.2 households (Table 3). We also observed increased transmissibility in BA.2 households from unvaccinated primary cases when compared to BA.1 households with an OR of 2.62 (95%-CI 1.96-3.52). The pattern of increased transmissibility in BA.2 households was not observed for fully vaccinated and booster-vaccinated primary cases, where the estimates were below 1 for BA.2 compared to BA.1 (Table 3).

**Table 3:**
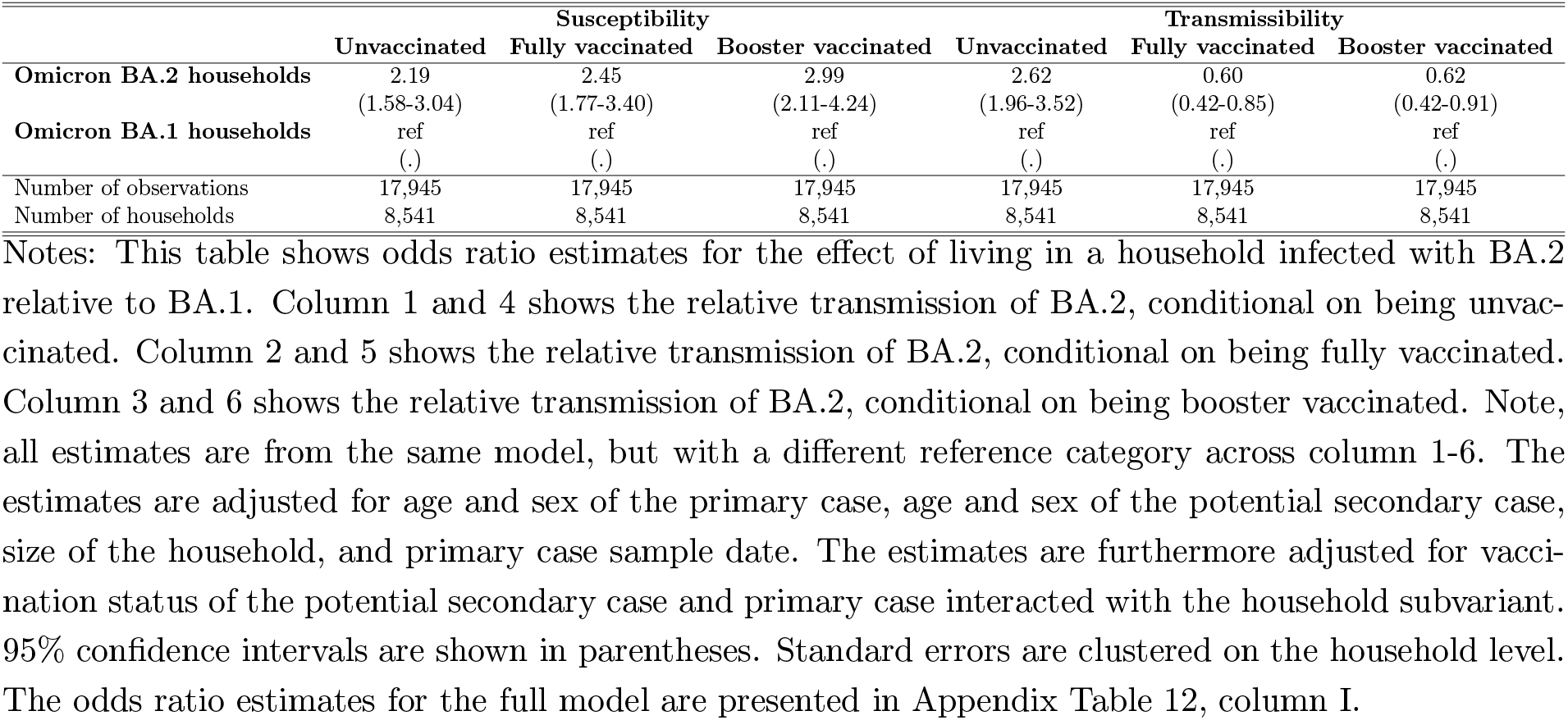
Relative effect of Omicron VOC BA.2 vs. BA.1

In the appendix, where we allowed for a 14-day follow-up, we found a 14-day SAR of 42% for BA.2 and 36% for BA.1 (appendix Figure 3).

The distribution of sample Ct values for unvaccinated primary cases showed that the viral load was overall higher for BA.2 cases than for BA.1 cases. This was not the case for fully vaccinated and booster-vaccinated individuals, where the distribution appeared to be the same (appendix Figure 5 and appendix Table 8).

## 5 Discussion

The present study shows that infection with the Omicron BA.2 subvariant generally lead to a higher SAR compared to BA.1 across all groups of sex, age, household sizes and immunity groups (Table 1). Furthermore, we found that booster-vaccinated individuals had a reduced susceptibility and transmissibility for both BA.1 and BA.2 compared to fully vaccinated individuals (Table 2). Efficient transmission to vaccinated individuals corroborates previous findings that the Omicron VOC possess immune evasive properties (Zhang et al., 2021; Lyngse et al., 2021b; Ferguson et al., 2021; Planas et al., 2021). However, both booster-vaccinated individuals and fully-vaccinated individuals had reduced susceptibility and transmissibility compared to unvaccinated individuals for both subvariants, suggesting that the effectiveness of vaccines remains significant (appendix Figure 6).

Both unvaccinated, fully vaccinated and booster-vaccinated individuals had a higher susceptibility for BA.2 compared to BA.1, indicating an inherent increased transmissibility of BA.2 (Table 3). However, the relative increase in susceptibility was significantly greater in vaccinated individuals compared to unvaccinated individuals (appendix Figure 6, which points towards immune evasive properties of the BA.2 conferring an even greater advantage for BA.2 in a highly vaccinated population such as Denmark. Because previous studies of the Omicron VOC has focused on the BA.1 (Pearson et al., 2021; Planas et al., 2021), new studies are needed to further investigate these properties for BA.2.

Unvaccinated individuals had a higher transmissibility with BA.2 compared to BA.1. Contrary to this, fully vaccinated and booster-vaccinated individuals had a reduced transmissibility, due to a significant negative interaction between subvariant and booster/fully vaccinated individuals compared to unvaccinated individuals (appendix Figure 6). This indicates that after a breakthrough infection, vaccination protects against further transmission, and more so for BA.2 than BA.1. This mechanism is only possible to identify in studies that take into account the exposure of individuals.

A potential mechanism for the higher transmissibility of unvaccinated individuals infected with BA.2 compared to BA.1 could be a higher viral load (appendix 7.3). No such difference was found for fully or booster-vaccinated individuals, which could be a result of a lower viral load in vaccinated individuals with a breakthrough infection (Puhach et al., 2022; Levine-Tiefenbrun et al., 2021; Lyngse et al., 2022).

The higher susceptibility and transmissibility among unvaccinated will likely result in even more extensive transmission of BA.2 in unvaccinated children in school settings and day care.

This study has a number of strengths. Firstly, Denmark is, to the best of our knowledge, the only country in the world that have been able to identify a large amount of both BA.1 and BA.2 cases in December 2021 and January 2022. Secondly, any bias introduced in the identification of the subvariants will presumably affect both BA.1 and BA.2 in a similar way. Third, this study draws on exhaustive population registers with a high quality of information covering the whole population. Fourth, potential secondary cases were frequently being tested: 84% one time, and 60-61% two times.

Some limitations apply to this study. The study period runs over Christmas 2021 and New Year’s Eve 2021/22, which are public holidays in Denmark. Despite government advice to limit social activity, it is likely that there has been considerable social mixing with family and friends outside the households during this period. Social mixing over the holidays in conjunction with the high incidence levels in Denmark during this period likely means that some secondary cases in this study are actually misclassified co-primary cases, i.e. infections picked up outside the household and testing positive after each other. However, this potential bias would be applicable to both subvariants. Moreover, our estimates were robust when only including primary cases from 5-11 January 2022 (appendix Table 12, model II) and when only including secondary cases found on day 2-7 or 3-7 (appendix Table 13, model VII and VIII).

The present household study showed a transmission advantage of Omicron BA.2 over BA.1. Although vaccinations, in particular booster vaccinations, did protect against infection, the 2.45 (fully vaccinated) and 2.99 (booster vaccinated) fold higher odds of infection in BA.2 households indicate that BA.2 as a phenotype represents a further step in immune evasion in the Omicron lineage. However, it is likely that this change came with an evolutionary cost for BA.2. To our surprise, we found a decreased transmissibility of BA.2 relative to BA.1 among fully vaccinated and booster vaccinated, with estimates of 0.60 and 0.62, respectively. Based on such a considerable loss in transmissibility among vaccinated individuals, it is not straightforward to predict the future trajectory of BA.2 relative to BA.1 or other potentially emerging variants.

Evolution of SARS-CoV-2 variants, including the Omicron VOC, is constantly evolving, especially during the current record high transmission in many countries. For public health, it is reassuring that BA.2, like BA.1, seems to be associated with favorable outcomes relative to the Delta variant, and that vaccines protect in particular against hospital admissions and severe illness (Wolter et al., 2022; Bager et al., 2022). Even with the emergence of BA.2, vaccines have an effect against infection, transmission and severe disease, although reduced compared to the ancestral variants. The combination of high incidence of a relative innocuous subvariant has raised optimism (Sundhedsministeriet, 2022). It is, however, important to follow the future evolution of the BA.2 subvariant closely, as well as future emergent subvariants. Thus, it is critical to maintain rapid high-quality WGS with random sampling as part of surveillance to continuously support the risk assessment of new variants, their impact on public health and to inform public health policy makers, when navigating during a pandemic.

## Acknowledgements

We thank Statens Serum Institut and The Danish Health Data Authority for collecting and providing access to data access. We also thank the rest of the Expert Group for Mathematical Modeling of COVID-19 at Statens Serum Institut for helpful discussions. The authors wish to thank the Danish Covid-19 Genome Consortium for genotyping SARS-CoV-2 positive samples. We thank Simon Kyllebæk Andersen (Department of Economics, University of Copenhagen) for proofreading the manuscript.

## Contributions

FPL performed all data analyses. MD calculated the contrasts between vaccination groups. FPL, CTK and LHM wrote the first draft. All other authors contributed to the discussion, revised the first draft and approved the submitted version.

## Supplementary Appendix

### 6 Background

This section provides some background characteristics for all of Denmark, i.e. not restricted to the study sample used for the analysis of household transmission.

#### 6.1 Number of tests

Table 4 shows the number of antigen (AG) tests and RT-PCR tests in Denmark from 1 December 2021 to 19 January 2022. The table also provides information on the number of successfully sequenced positive RT-PCR tests by SARS-CoV-2 variant, including their relative proportion. On 20 December 2021, Omicron BA.2 comprised 5% of all infections, and Omicron BA.1 comprised 64%, while Delta comprised 30%. By 11 January 2022, the proportions were 47%, 53%, and 0%, respectively.

**Table 4:**
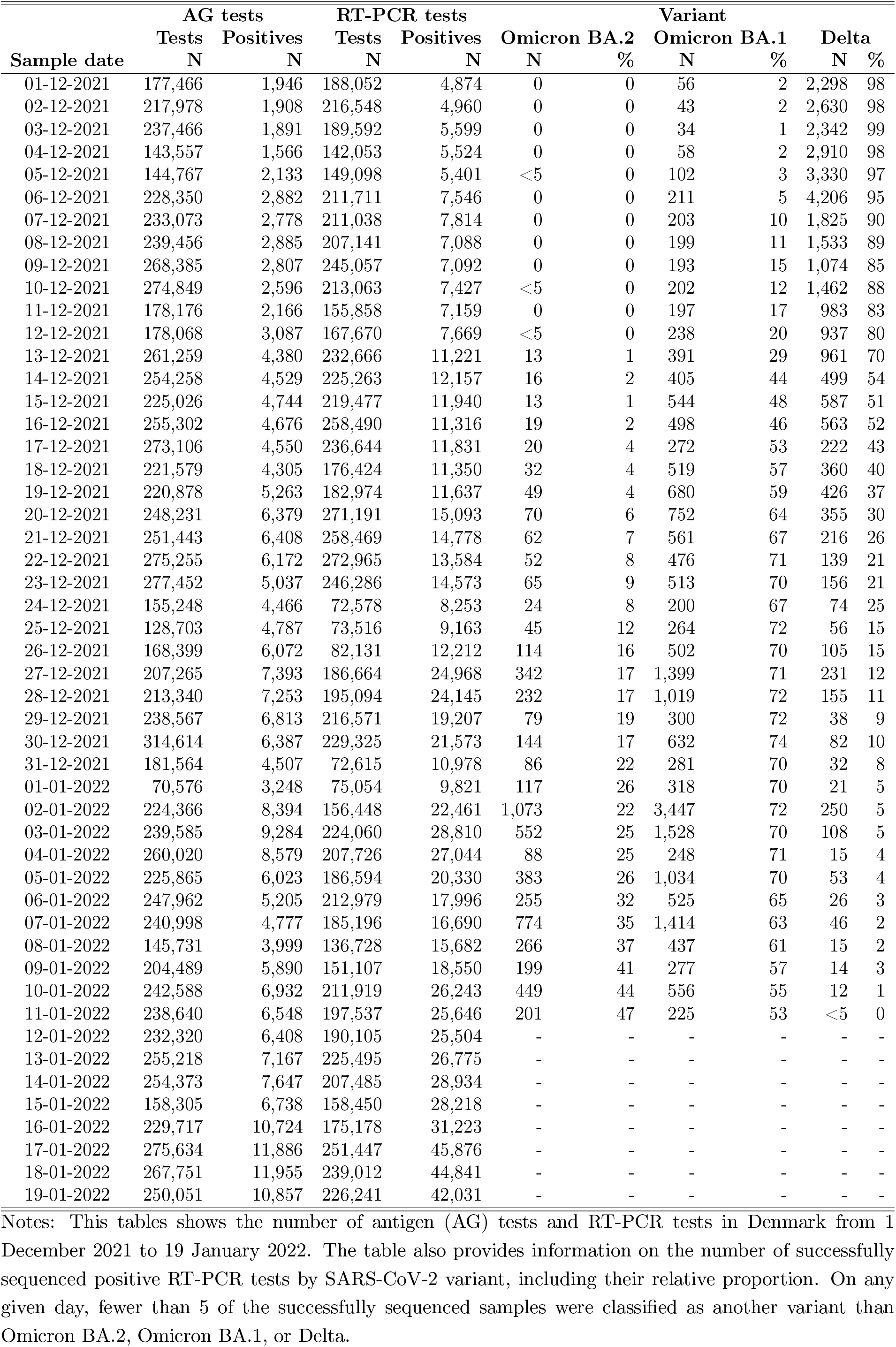
Number of tests in Denmark, 1 December 2021–19 January 2022

**Table 5:**
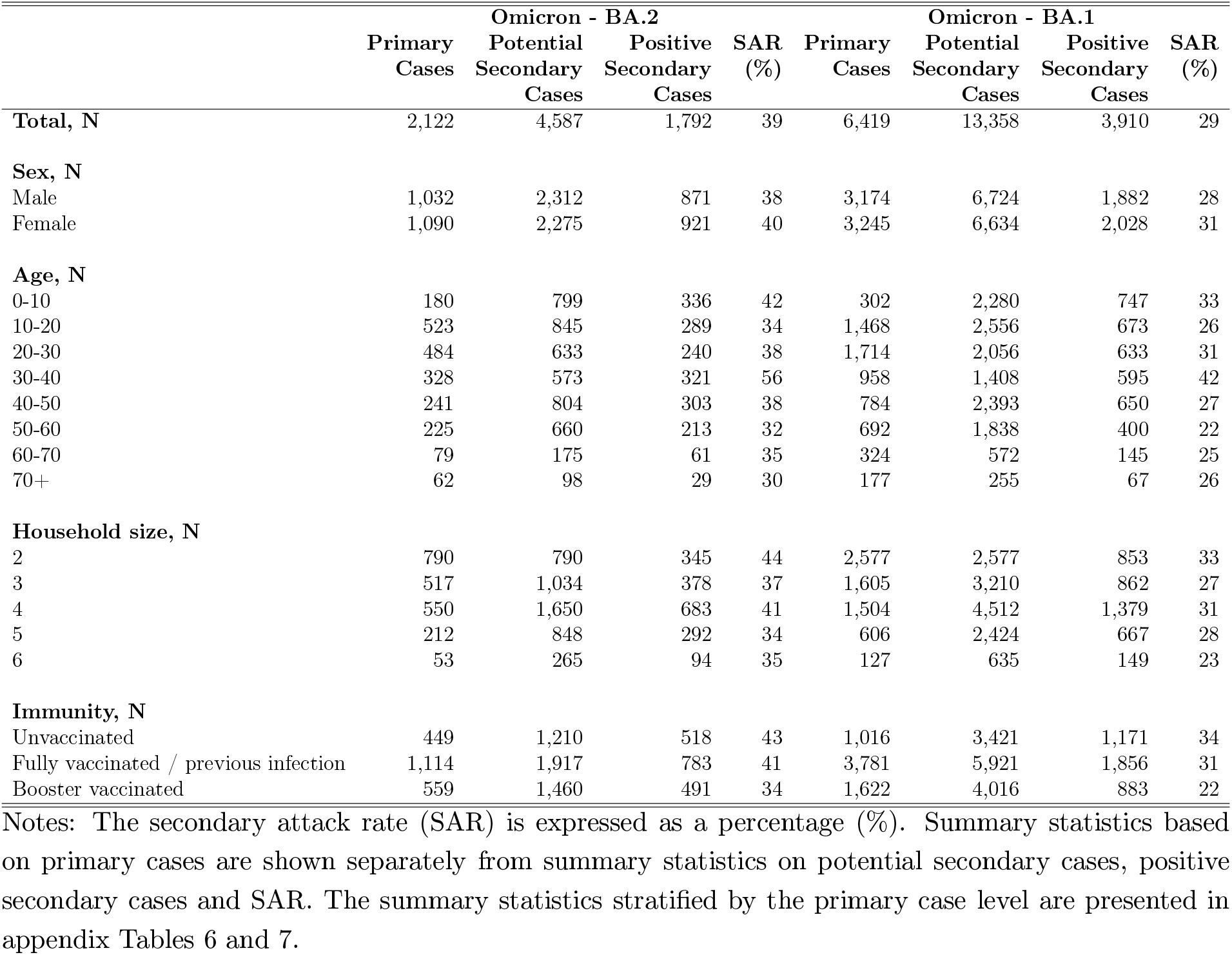
Summary Statistics

**Table 6:**
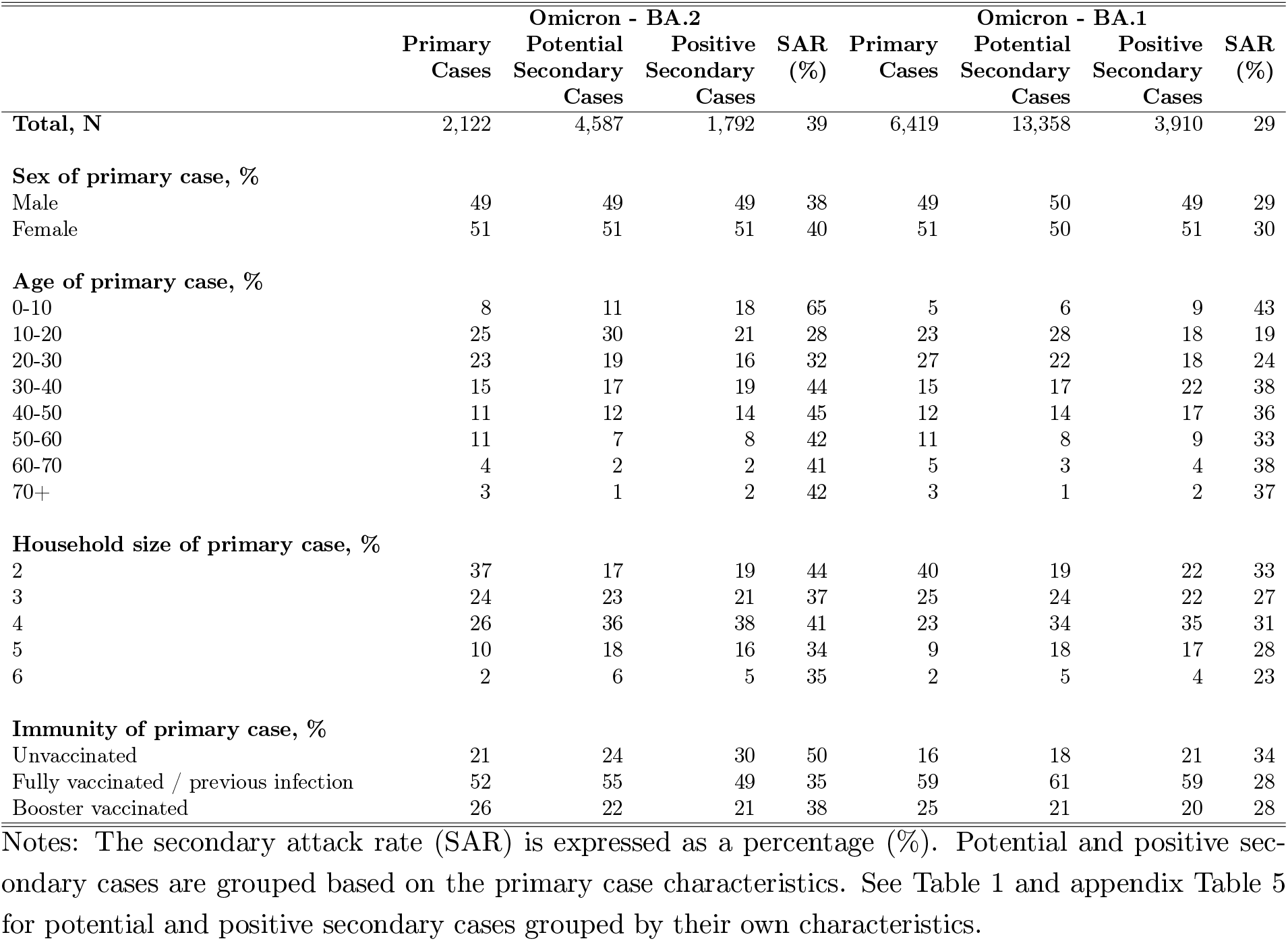
Summary Statistics, stratified by primary case level

|  | Omicron - BA.2 |  |  |  | Omicron - BA.1 |  |  |  |
| --- | --- | --- | --- | --- | --- | --- | --- | --- |
|  | Primary<br>Cases | Potential<br>Secondary<br>Cases | Positive<br>Secondary<br>Cases | SAR<br>(%) | Primary<br>Cases | Potential<br>Secondary<br>Cases | Positive<br>Secondary<br>Cases | SAR<br>(%) |
| **Total, N** | 2,122 | 4,587 | 1,792 | 39 | 6,419 | 13,358 | 3,910 | 29 |
| **Sex of primary case, %** |  |  |  |  |  |  |  |  |
| Male | 49 | 49 | 49 | 38 | 49 | 50 | 49 | 29 |
| Female | 51 | 51 | 51 | 40 | 51 | 50 | 51 | 30 |
| **Age of primary case, %** |  |  |  |  |  |  |  |  |
| 0-10 | 8 | 11 | 18 | 65 | 5 | 6 | 9 | 43 |
| 10-20 | 25 | 30 | 21 | 28 | 23 | 28 | 18 | 19 |
| 20-30 | 23 | 19 | 16 | 32 | 27 | 22 | 18 | 24 |
| 30-40 | 15 | 17 | 19 | 44 | 15 | 17 | 22 | 38 |
| 40-50 | 11 | 12 | 14 | 45 | 12 | 14 | 17 | 36 |
| 50-60 | 11 | 7 | 8 | 42 | 11 | 8 | 9 | 33 |
| 60-70 | 4 | 2 | 2 | 41 | 5 | 3 | 4 | 38 |
| 70+ | 3 | 1 | 2 | 42 | 3 | 1 | 2 | 37 |
| **Household size of primary case, %** |  |  |  |  |  |  |  |  |
| 2 | 37 | 17 | 19 | 44 | 40 | 19 | 22 | 33 |
| 3 | 24 | 23 | 21 | 37 | 25 | 24 | 22 | 27 |
| 4 | 26 | 36 | 38 | 41 | 23 | 34 | 35 | 31 |
| 5 | 10 | 18 | 16 | 34 | 9 | 18 | 17 | 28 |
| 6 | 2 | 6 | 5 | 35 | 2 | 5 | 4 | 23 |
| **Immunity of primary case, %** |  |  |  |  |  |  |  |  |
| Unvaccinated | 21 | 24 | 30 | 50 | 16 | 18 | 21 | 34 |
| Fully vaccinated / previous infection | 52 | 55 | 49 | 35 | 59 | 61 | 59 | 28 |
| Booster vaccinated | 26 | 22 | 21 | 38 | 25 | 21 | 20 | 28 |

Notes: The secondary attack rate (SAR) is expressed as a percentage (%). Potential and positive secondary cases are grouped based on the primary case characteristics. See Table 1 and appendix Table 5 for potential and positive secondary cases grouped by their own characteristics.

**Table 7:** Summary Statistics, stratified by primary case level

|  | Omicron - BA.2 |  |  |  | Omicron - BA.1 |  |  |  |
| --- | --- | --- | --- | --- | --- | --- | --- | --- |
|  | Primary Cases | Potential Secondary Cases | Positive Secondary Cases | SAR (%) | Primary Cases | Potential Secondary Cases | Positive Secondary Cases | SAR (%) |
| **Total, N** | 2,122 | 4,587 | 1,792 | 39 | 6,419 | 13,358 | 3,910 | 29 |
| **Sex of primary case, N** |  |  |  |  |  |  |  |  |
| Male | 1,032 | 2,270 | 873 | 38 | 3,174 | 6,643 | 1,928 | 29 |
| Female | 1,090 | 2,317 | 919 | 40 | 3,245 | 6,715 | 1,982 | 30 |
| **Age of primary case, N** |  |  |  |  |  |  |  |  |
| 0-10 | 180 | 495 | 324 | 65 | 302 | 828 | 353 | 43 |
| 10-20 | 523 | 1,360 | 375 | 28 | 1,468 | 3,782 | 720 | 19 |
| 20-30 | 484 | 873 | 278 | 32 | 1,714 | 2,903 | 707 | 24 |
| 30-40 | 328 | 783 | 348 | 44 | 958 | 2,242 | 860 | 38 |
| 40-50 | 241 | 572 | 256 | 45 | 784 | 1,886 | 681 | 36 |
| 50-60 | 225 | 342 | 144 | 42 | 692 | 1,134 | 369 | 33 |
| 60-70 | 79 | 95 | 39 | 41 | 324 | 391 | 149 | 38 |
| 70+ | 62 | 67 | 28 | 42 | 177 | 192 | 71 | 37 |
| **Household size of primary case, N** |  |  |  |  |  |  |  |  |
| 2 | 790 | 790 | 345 | 44 | 2,577 | 2,577 | 853 | 33 |
| 3 | 517 | 1,034 | 378 | 37 | 1,605 | 3,210 | 862 | 27 |
| 4 | 550 | 1,650 | 683 | 41 | 1,504 | 4,512 | 1,379 | 31 |
| 5 | 212 | 848 | 292 | 34 | 606 | 2,424 | 667 | 28 |
| 6 | 53 | 265 | 94 | 35 | 127 | 635 | 149 | 23 |
| **Immunity of primary case, N** |  |  |  |  |  |  |  |  |
| Unvaccinated | 449 | 1,083 | 543 | 50 | 1,016 | 2,417 | 816 | 34 |
| Fully vaccinated / previous infection | 1,114 | 2,507 | 875 | 35 | 3,781 | 8,152 | 2,315 | 28 |
| Booster vaccinated | 559 | 997 | 374 | 38 | 1,622 | 2,789 | 779 | 28 |

Notes: The secondary attack rate (SAR) is expressed as a percentage (%). Potential and positive secondary cases are grouped based on the primary case characteristics. See Table 1 and appendix Table 5 for potential and positive secondary cases grouped by their own characteristics.

#### 6.2 Sample selection to WGS

In Denmark, individuals can be tested in the community track (TestCenter Denmark) or in the healthcare track (Hospitals), which includes hospitalized patients, nursing home residents, and healthcare personnel (see Schønning et al. (2021) for elaboration). Only a proportion of all positive RT-PCR tests were sampled for whole genome sequencing (WGS). Both TestCenter Denmark and Hospitals sample positive RT-PCR tests randomly for WGS. However, all hospitalized patients were tested for SARS-CoV-2 and all positive tests were subject to WGS for treatment purposes. Figure 2 shows the sampling probability for WGS for within the study period by TestCenter Denmark and Hospitals. Panel a and b shows the sampling probability by age. For positive RT-PCR tests at TestCenter Denmark, there was no selection bias on age, whereas in hospitals, there was an increased sampling probability by age for young children and elderly. Panel c shows the sampling probability for WGS by sample Ct value for TestCenter Denmark (we only obtained Ct values from TestCenter Denmark). There was no sampling bias for Ct values <35. The probability of obtaining a successfully sequenced genome was correlated with the sample Ct value.

**Figure 2:**
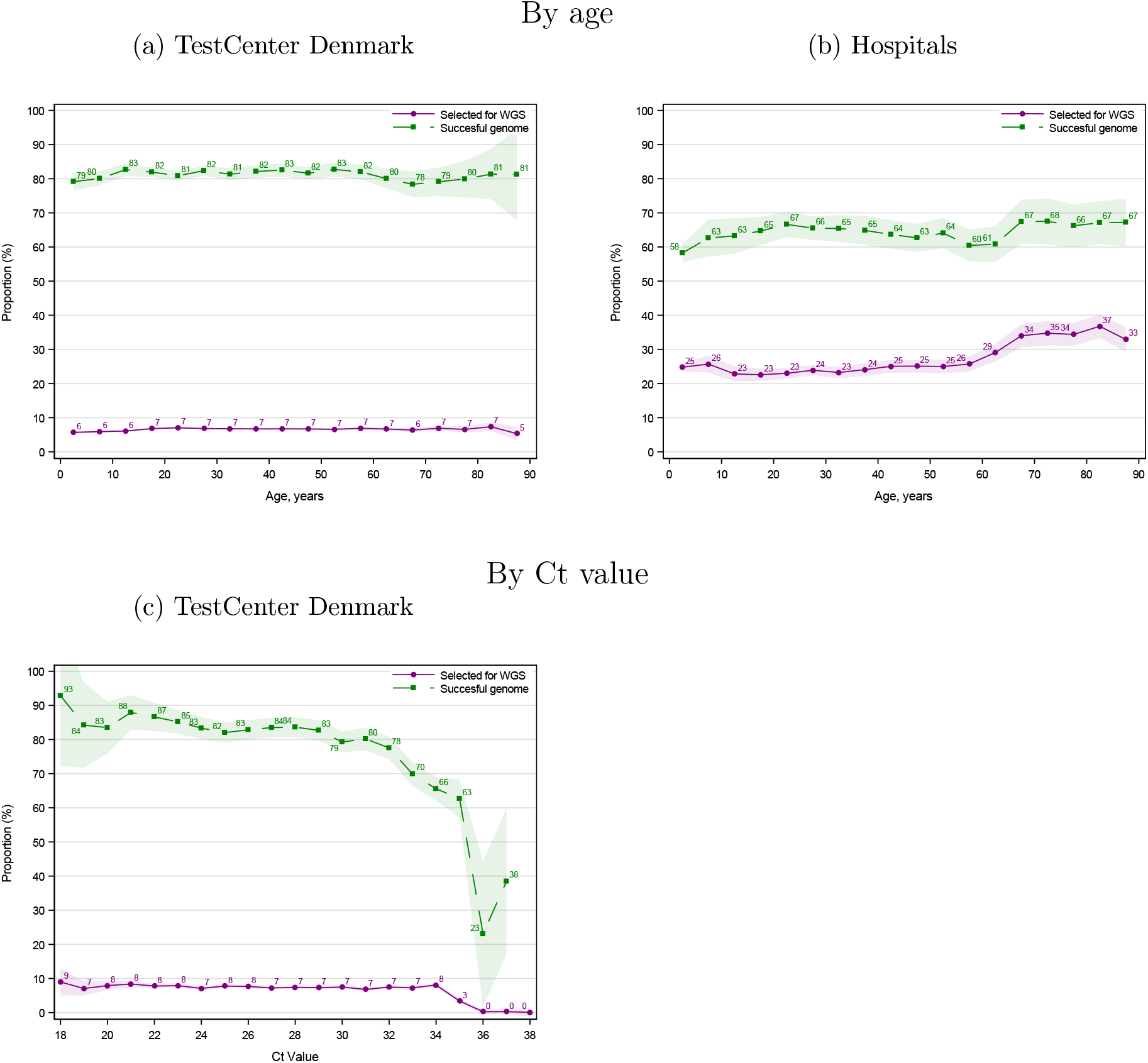
WGS sampling probability of positive RT-PCR tests By age Notes: This figure shows the sampling probability of positive RT-PCR tests for WGS by testing place (TestCenter Denmark and Hospitals). Only Ct values from TestCenter Denmark were available. The shaded areas show the 95% confidence bands clustered on the household level.

### 7 Descriptive analyses

#### 7.1 Summary statistics

In this section, we present additional summary statistics.

#### 7.2 Testing dynamics, 14-day follow-up

In this section, we present evidence of the testing dynamics over a 14-day follow-up period. Figure 3 presents the probability of being tested and testing positive over a 14-day followup period instead of a 7-day follow-up period, as used in Figure 1. Figure 4 presents the SAR for households infected with the Omicron BA.1, BA.2, and Delta VOC, as well as those without an identified variant.

**Figure 3:**
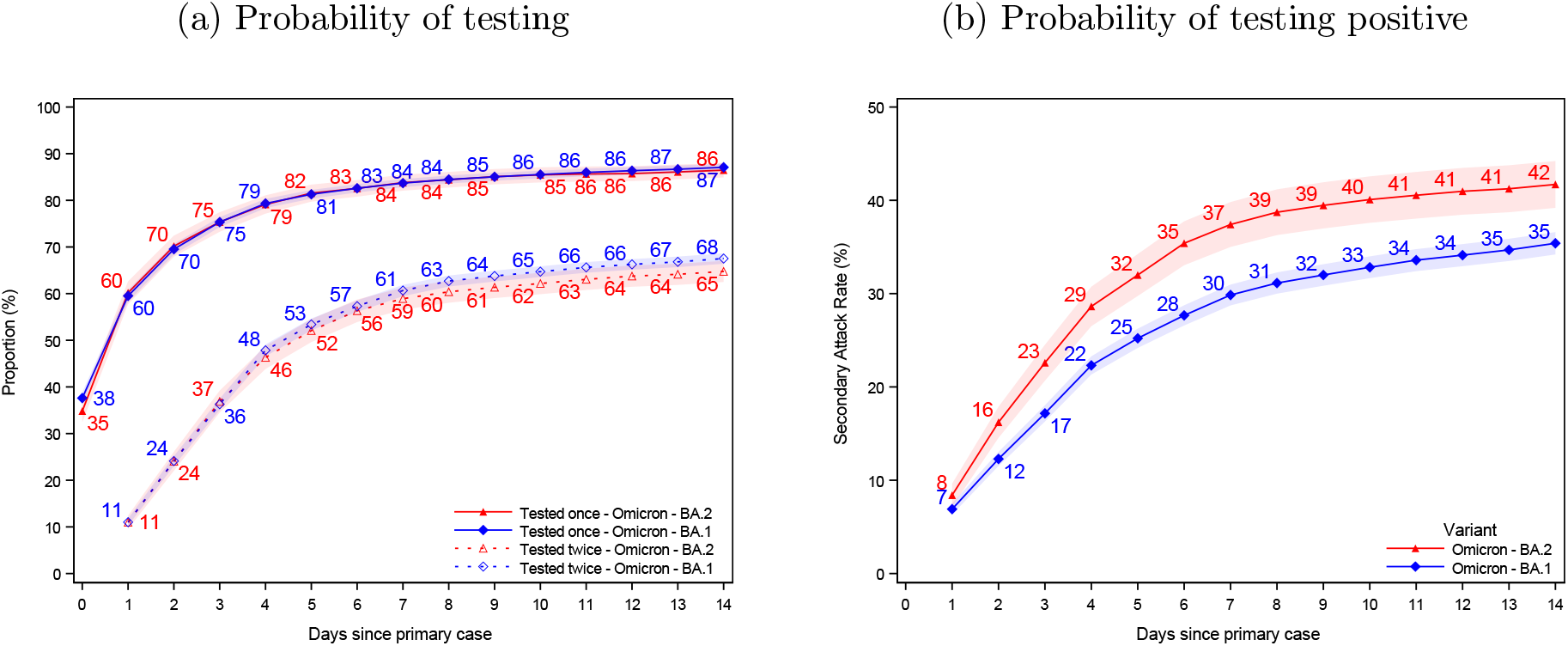
Probability of being tested and testing positive, 14-day follow-up Notes: Panel (a) shows the probability of potential secondary cases being tested after a primary case has been identified within the household. Panel (b) shows the probability of potential secondary cases that test positive subsequently to a primary case being identified within the household. Note that the latter is not conditional on being tested, i.e. the denominator contains test negative individuals and untested individuals. The x-axes shows the days since the primary case tested positive, and the y-axes shows the proportion of individuals either being tested (a) or testing positive (b) with either antigen or RT-PCR tests, based on the subvariant of the primary case. The SAR for each day relative to the primary case can be read directly from panel (b). For example, the SAR on day 7 is 37% for BA.2 (red) and 30% for BA.1 (blue), whereas the SAR on day 14 is 42% and 35%, respectively. The shaded areas show the 95% confidence bands clustered on the household level. To allow for a 14-day follow-up, only primary cases with samples from 20 December 2021 to 5 January 2022 were included in this figure. Appendix Figure 4 also presents the 14-day SAR for the Delta VOC and those without a known variant.

**Figure 4:**
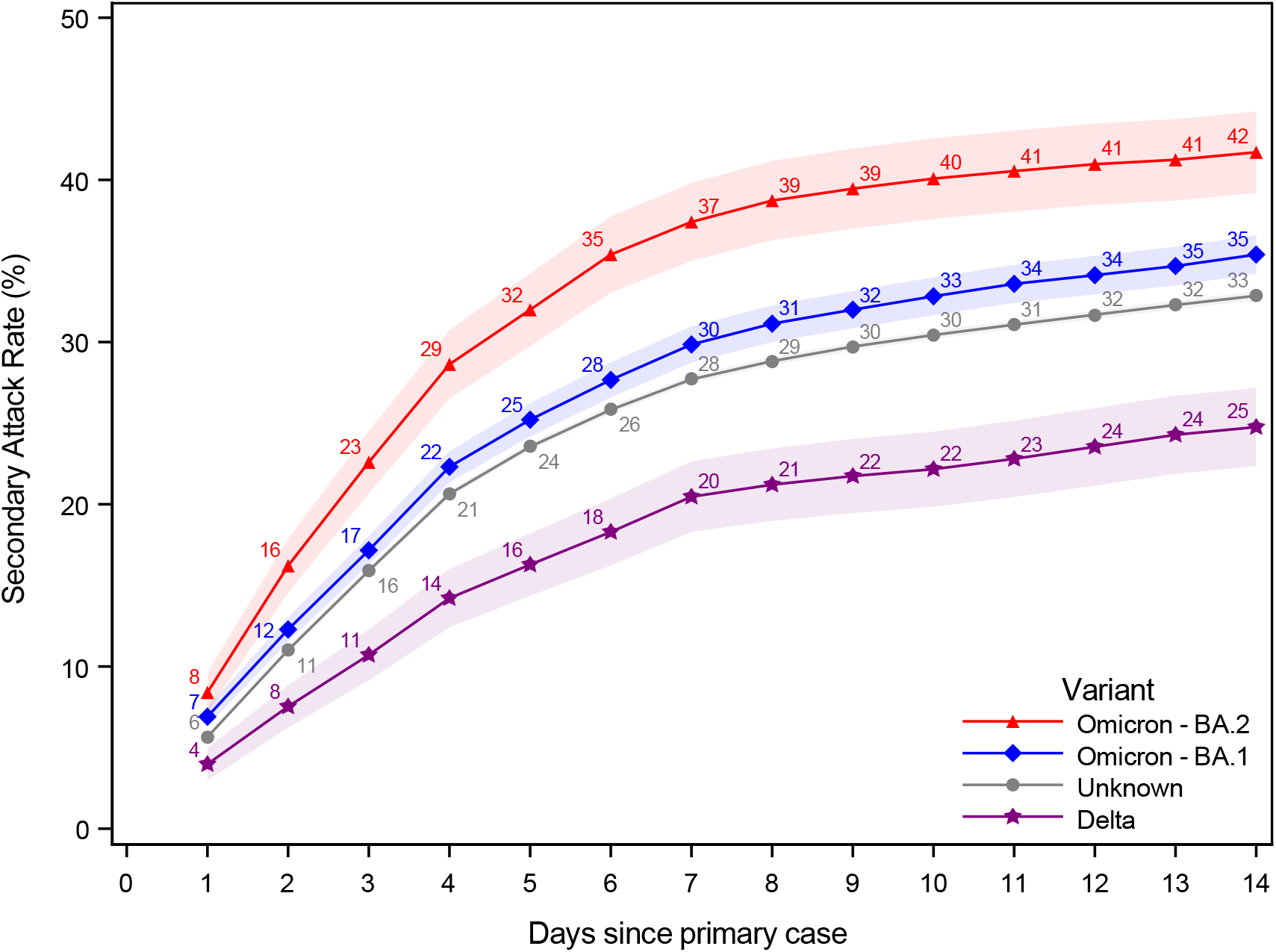
Probability of testing positive, 14-day follow-up Notes: This figure shows the probability of potential secondary cases that test positive subsequently to a primary case being identified within the household in a 14-day follow-up period. Note that the latter is not conditional on being tested, i.e. the denominator contains test negative individuals and untested individuals. The x-axes shows the days since the primary case tested positive, and the y-axes shows the proportion of individuals testing positive with either antigen or RT-PCR tests, based on the subvariant of the primary case. The SAR for each day relative to the primary case can be read directly from the figure. For example, the SAR on day 14 is 42% for Omicron BA.2, 35% for BA.1, 33% for those without a known variant, and 25% for Delta. The shaded areas show the 95% confidence bands clustered on the household level. To allow for a 14-day follow-up, only primary cases with samples from 20 December 2021 to 5 January 2022 were included in this figure.

#### 7.3 Viral load of primary cases

This section provides descriptive statistics on the viral load of the primary case samples. Figure 5 shows the density plots of sample Ct values for primary cases infected with Omicron BA.1 and BA.2 stratified by their vaccination status. The distributional values are presented in Table 8. In particular, unvaccinated primary cases infected with BA.2 have a higher sample viral load (lower Ct value), with the median primary case having a 1.6 point lower sample Ct value, corresponding to 0.4 of a standard deviation.

**Figure 5:**
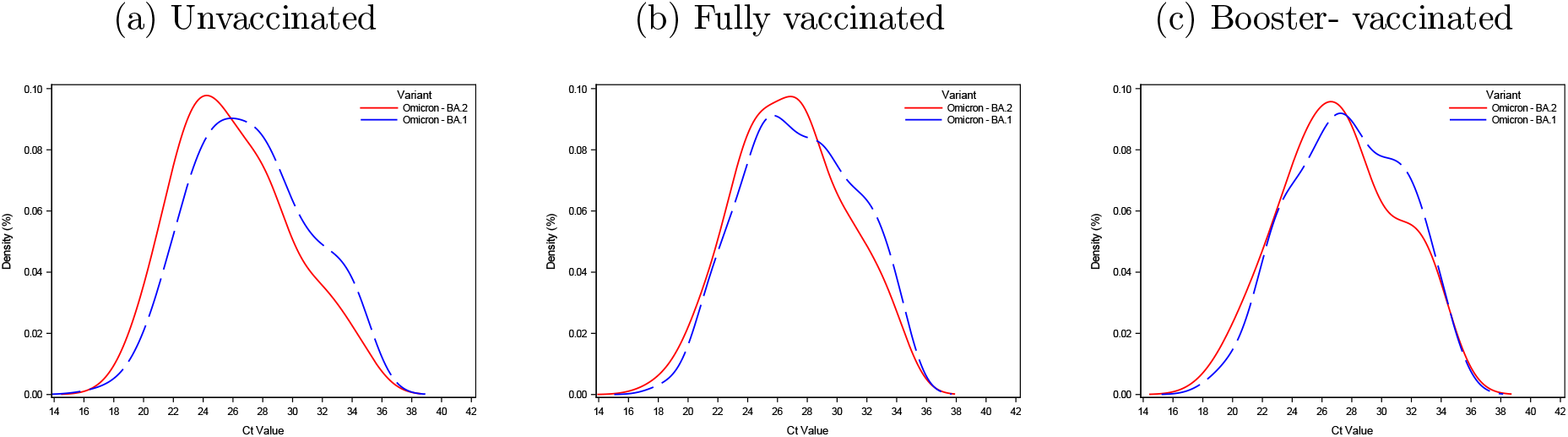
Ct values of primary cases Notes: This figure shows the density plots for primary cases infected with Omicron BA.1 and BA.2 stratified by their vaccination status.

**Table 8:** Ct values of primary cases

| Vaccination status | Subvariant | Q1 | Median | Q3 | Mean | STD | N |
| --- | --- | --- | --- | --- | --- | --- | --- |
| Booster vaccinated | Omicron - BA.1 | 25.01 | 27.65 | 30.80 | 27.72 | 3.71 | 1,622 |
|  | Omicron - BA.2 | 24.48 | 27.09 | 30.06 | 27.20 | 3.82 | 559 |
|  | Difference / STD | -0.14 | -0.14 | -0.19 | -0.13 |  |  |
| Fully vaccinated | Omicron - BA.1 | 24.65 | 27.33 | 30.48 | 27.50 | 3.76 | 3,781 |
|  | Omicron - BA.2 | 24.10 | 26.97 | 29.47 | 26.88 | 3.68 | 1,114 |
|  | Difference / STD | -0.15 | -0.09 | -0.26 | -0.16 |  |  |
| Not vaccinated | Omicron - BA.1 | 24.14 | 27.02 | 29.97 | 27.20 | 3.91 | 1,016 |
|  | Omicron - BA.2 | 23.20 | 25.42 | 28.55 | 26.07 | 3.80 | 449 |
|  | Difference / STD | -0.24 | -0.41 | -0.36 | -0.29 |  |  |

Notes: This table provides distributional values for the Ct values of primary case samples. "Difference / STD" denotes the difference of primary cases with BA.2 and BA.1 relative to the standard deviation of BA.1 primary cases, within vaccination group.

### 8 Alternative presentation of main results

#### 8.1 Contrasts

In this section, we present some of our main estimates in an alternative way, showing the estimates for comparison of different vaccination groups.

Figure 6 shows a full comparison of our main estimates across vaccination groups with different reference groups. We can see the relative effect of vaccination dependent on their vaccination status by choosing the *Contrast* (column) and compare their *Transmissibility* to the vaccination status of a similar primary case by choosing the *Reference* (row). For example, unvaccinated primary cases (Contrast=unvaccinated) compared to fully vaccinated primary cases (Reference=fully vaccinated) have an increased transmissibility of OR=1.13 when infected with BA.1 (blue) and OR=1.23 when infected with BA.2 (red). The interaction term of BA.2 on transmissibility has an OR=1.09 (black). This interaction term can be interpreted as the additional OR associated with BA.2 (relative to BA.1) within the comparison. Similarly, we can see the relative transmissibility for unvaccinated primary cases (Contrast=unvaccinated) compared to booster-vaccinated primary cases (Reference=booster). Primary cases with BA.1 have an OR=1.36 for transmissibility and primary cases with BA.2 have an OR=1.53 for transmissibility. The interaction term is OR=1.12. The estimates for *Susceptibility* is read in a similar way, but for potential secondary cases. Lastly, the *Combined* effect of vaccination shows the effect of both the primary case and potential secondary case having the same vaccination status. Thus, for households infected with BA.1, the OR=1.40 if both the primary case and potential secondary case are unvaccinated (Contrast=Unvaccinated) compared to when both are fully vaccinated (Reference=Fully vaccinated). Note, the model estimates for reference=Fully vaccinated are found in Table 10.

**Figure 6:**
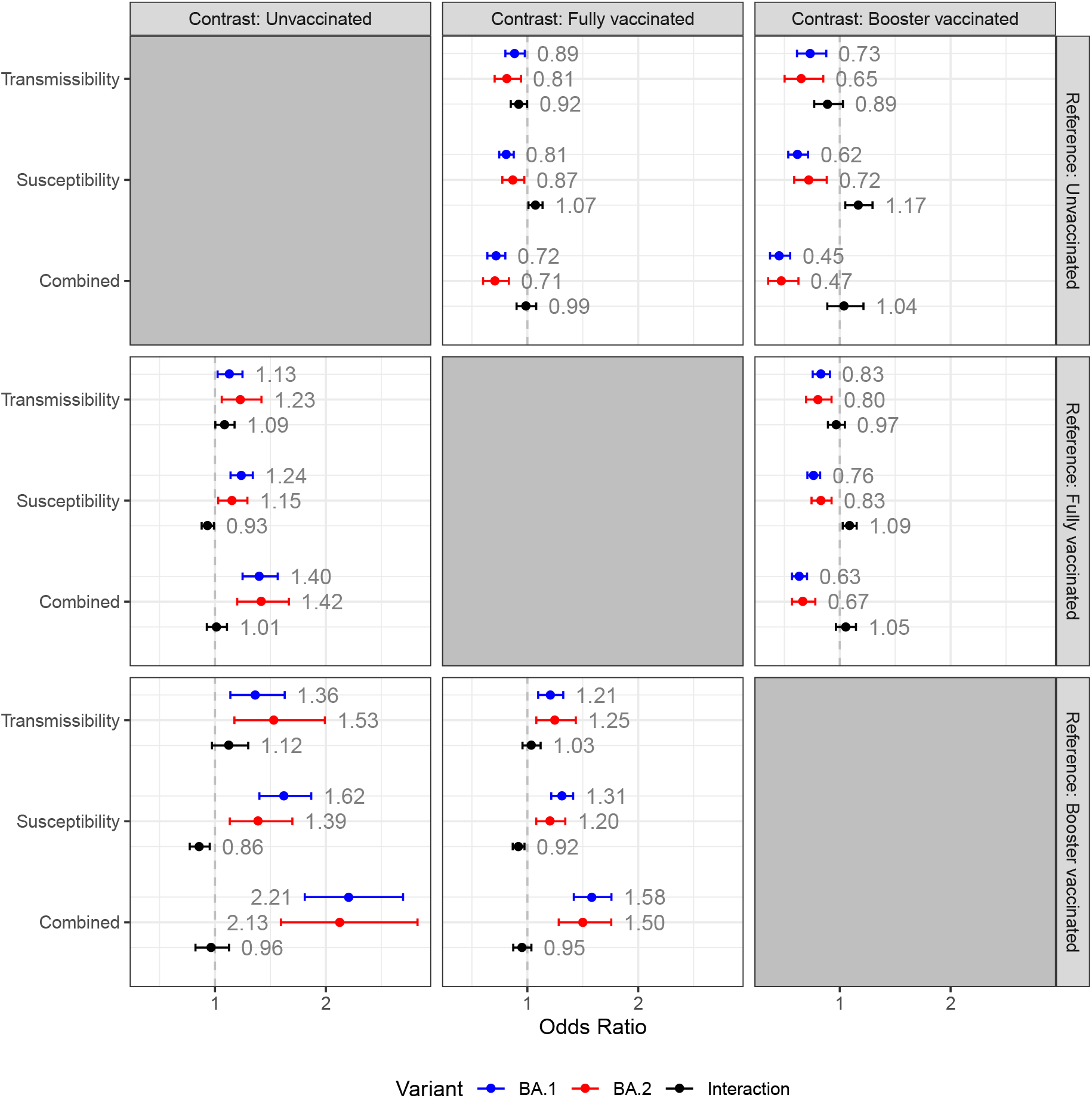
Effect of vaccination, contrast plot Notes: This figure shows a full comparison of our main estimates across vaccination groups with different reference groups. We can see the relative effect of vaccination dependent on their vaccination status by choosing the *Contrast* (column) and compare their *Transmissibility* to the vaccination status of a similar primary case by choosing the *Reference* (row). For example, unvaccinated primary cases (Contrast=unvaccinated) compared to fully vaccinated primary cases (Reference=fully vaccinated) have an increased transmissibility of OR=1.13 when infected with BA.1 (blue) and OR=1.23 when infected with BA.2 (red). The interaction term of BA.2 on transmissibility has an OR=1.09 (black). This interaction term can be interpreted as the additional OR associated with BA.2 (relative to BA.1) within the comparison. Note that the top/right subplots are simply the inverse of the lower/left subplots. 95%-confidence intervals. Standard errors are clustered on the household level.

Table 9 shows the OR for infection with BA.2 compared to BA.1 for each combination of vaccination status of both the primary case and potential secondary case. The relative transmission of BA.2 compared to BA.1 is higher across all combinations of vaccination groups. The relative transmission of BA.2 compared to BA.1 is generally higher for unvaccinated primary cases across all vaccination groups of potential secondary cases. Table 10 shows the model estimates (similar to Table 12, model I) with interaction terms instead of a full specification of contrasts.

**Table 9:**
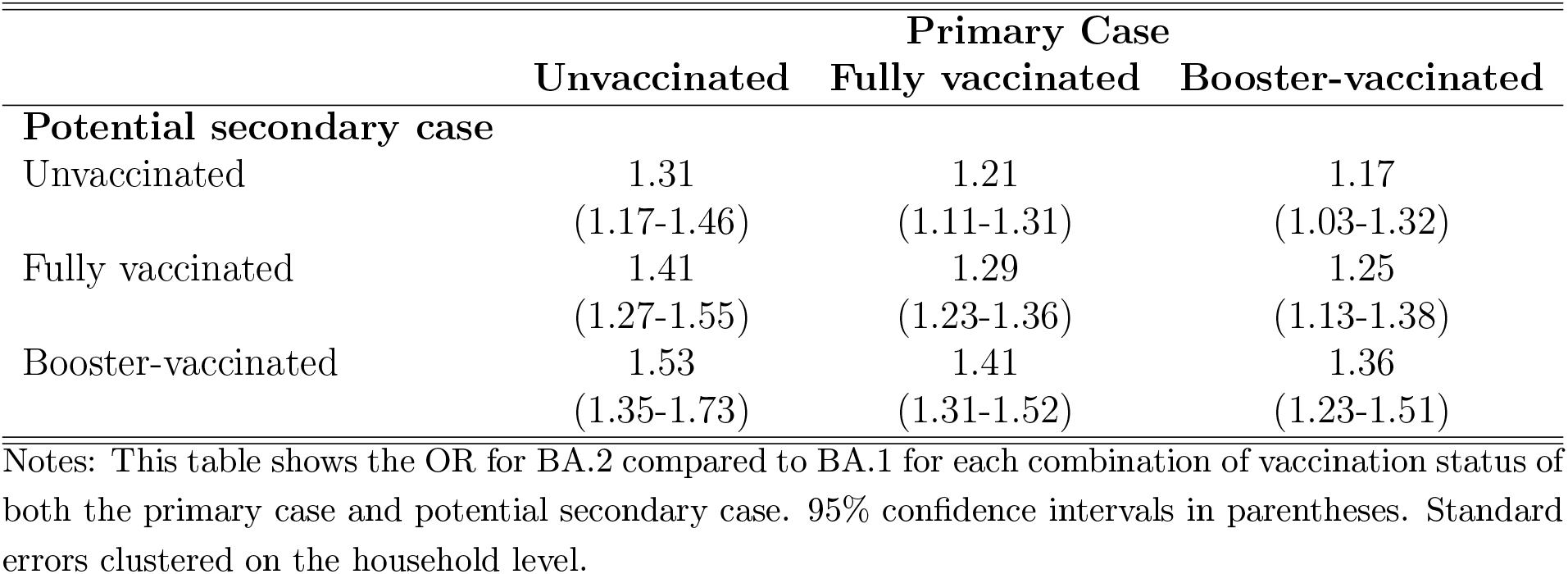
OR estimates of BA.2 compared to BA.1 by vaccination status

**Table 10:** Model estimates

|  | Estimate | (95%-CI) |
| --- | --- | --- |
| Intercept | -0.58 | (-0.68;-0.48) |
| Omicron BA.2 | 0.26 | (0.21;0.31) |
| **Potential secondary case, vaccination status** |  |  |
| Booster-vaccinated | -0.27 | (-0.35;-0.19) |
| Fully vaccinated | ref | (.) |
| Not-vaccinated | 0.21 | (0.13;0.29) |
| **Primary case, vaccination status** |  |  |
| Booster-vaccinated | -0.19 | (-0.28;-0.09) |
| Fully vaccinated | ref | (.) |
| Not-vaccinated | 0.12 | (0.02;0.22) |
| **Potential secondary case, vaccination status** |  |  |
| Omicron BA.2 X Booster-vaccinated | 0.08 | (0.03;0.14) |
| Omicron BA.2 X Fully vaccinated | ref | (.) |
| Omicron BA.2 X Not-vaccinated | -0.07 | (-0.13;-0.01) |
| **Primary case, vaccination status** |  |  |
| Omicron BA.2 X Booster-vaccinated | -0.03 | (-0.11;0.04) |
| Omicron BA.2 X Fully vaccinated | ref | (.) |
| Omicron BA.2 X Not-vaccinated | 0.08 | (0.01;0.16) |
| **Primary case age** |  |  |
| 0-10 | 0.30 | (0.12;0.49) |
| 10-20 | -0.76 | (-0.88;-0.64) |
| 20-30 | ref | (.) |
| 30-40 | -0.06 | (-0.18;0.06) |
| 40-50 | 0.09 | (-0.03;0.21) |
| 50-60 | 0.03 | (-0.10;0.17) |
| 60-70 | 0.38 | (0.17;0.59) |
| 70+ | 0.71 | (0.38;1.04) |
| **Potential secondary case age** |  |  |
| 0-10 | -0.11 | (-0.23;0.02) |
| 10-20 | -0.12 | (-0.22;-0.02) |
| 20-30 | ref | (.) |
| 30-40 | 0.48 | (0.37;0.58) |
| 40-50 | 0.26 | (0.16;0.36) |
| 50-60 | 0.00 | (-0.11;0.11) |
| 60-70 | -0.18 | (-0.35;-0.01) |
| 70+ | -0.56 | (-0.84;-0.27) |
| **Household size** |  |  |
| 2 | 0.27 | (0.18;0.36) |
| 3 | -0.02 | (-0.11;0.06) |
| 4 | 0.07 | (-0.01;0.15) |
| 5 | -0.08 | (-0.19;0.03) |
| 6 | ref | (.) |
| **Primary case sex** |  |  |
| Male | ref | (.) |
| Female | 0.14 | (0.07;0.20) |
| **Potential secondary case sex** |  |  |
| Male | ref | (.) |
| Female | 0.06 | (-0.02;0.15) |
| **Primary case sample date** |  |  |
| 20DEC2021 | -0.15 | (-0.34;0.05) |
| 21DEC2021 | 0.02 | (-0.21;0.24) |
| 22DEC2021 | 0.15 | (-0.08;0.39) |
| 23DEC2021 | 0.09 | (-0.17;0.34) |
| 24DEC2021 | 0.20 | (-0.20;0.59) |
| 25DEC2021 | 0.46 | (0.12;0.80) |
| 26DEC2021 | 0.20 | (-0.08;0.47) |
| 27DEC2021 | 0.06 | (-0.10;0.23) |
| 28DEC2021 | -0.01 | (-0.19;0.18) |
| 29DEC2021 | 0.14 | (-0.16;0.44) |
| 30DEC2021 | -0.04 | (-0.26;0.18) |
| 31DEC2021 | ref | (.) |
| 01JAN2022 | -0.03 | (-0.32;0.25) |
| 02JAN2022 | -0.05 | (-0.16;0.05) |
| 03JAN2022 | -0.09 | (-0.23;0.05) |
| 04JAN2022 | -0.16 | (-0.48;0.15) |
| 05JAN2022 | -0.36 | (-0.54;-0.19) |
| 06JAN2022 | -0.21 | (-0.43;0.00) |
| 07JAN2022 | -0.15 | (-0.29;-0.01) |
| 08JAN2022 | -0.09 | (-0.32;0.14) |
| 09JAN2022 | 0.00 | (-0.24;0.24) |
| 10JAN2022 | -0.04 | (-0.21;0.14) |
| 11JAN2022 | 0.02 | (-0.26;0.31) |
| Number of observations | 17,945 |  |
| Number of households | 8,541 |  |

Notes: This table provides model estimates of our main specification (model 1) with an interaction effect. 95% confidence intervals are shown in parentheses. Standard errors are clustered on the household level.

**Table 11:** Model selection

| Model | I | II | III | IV |
| --- | --- | --- | --- | --- |
| AIC | 21,245 | 21,249 | 21,260 | 21,263 |
| Omicron BA.2 household | YES | YES | YES | YES |
| Potential secondary case vaccination status | YES | YES | YES | YES |
| Primary case vaccination status | YES | YES | YES | YES |
| Potential secondary case vaccination status X Omicron BA.2 household | YES | YES | NO | NO |
| Primary case vaccination status X Omicron BA.2 household | YES | NO | YES | NO |
| Primary case age | YES | YES | YES | YES |
| Potential secondary case age | YES | YES | YES | YES |
| Household size | YES | YES | YES | YES |
| Primary case sex | YES | YES | YES | YES |
| Potential secondary case sex | YES | YES | YES | YES |
| Number of observations | 17,945 | 17,945 | 17,945 | 17,945 |
| Number of households | 8,541 | 8,541 | 8,541 | 8,541 |

Notes: This table provides estimates of the goodness of fit for the model. Model I includes an interaction with Omicron BA.2 both for susceptibility and transmissibility, and is the one used in the study. Model II includes an interaction with Omicron BA.2 only for susceptibility. Model III includes an interaction with Omicron BA.2 only for transmissibility. Model IV includes neither an interaction with Omicron BA.2 only for susceptibility nor transmissibility. AIC = Akaike information criterion.

##### Method for estimating the constrast

To estimate the contrast, we used the estimates and variance-covariance matrix from the fitted model (Table 10) to generate 10,000 correlated Monte Carlo estimates of the model parameters. These estimates were then used to calculate log odds ratios representing the total effect of BA.2 compared to BA.1 for all possible pairwise contrasts within vaccination groups. This was done for transmissibility and susceptibility, as well as a combined effect calculated as the sum of these. In addition, the interaction between subvariant and vaccination group was also calculated for each pairwise contrast within vaccination groups. Results were summarised as mean and 95%-confidence intervals of the estimates, before exponentiation for interpretation as odds ratios (Figure 6). These analyses were performed in R version 4.1.2 (R Core Team, 2021).

### 9 Robustness

#### 9.1 Model selection

#### 9.2 Intra-household correlation of variants

An obvious concern in transmission studies is the linkage of primary cases to their potential secondary cases and positive secondary cases. In previous studies we have used the same method as in the current study (Lyngse et al., 2021a,b). In those studies, we investigated the household transmission between different SARS-CoV-2 variants and found an intrahousehold correlation of variants (i.e. the probability that a positive secondary case was infected with the same variant as the primary case) of 96-99%. In the present study, we are limited by only having a low number of positive secondary cases with a successfully sequenced genome. This is due to both the laboratory time needed from a positive test result to having a successfully sequenced genome from the sample and a relatively low sampling probability for WGS. Of all the 5,702 positive secondary samples in the current study, only 23 of them had a successfully sequenced genome at the time of analysis. All of these 23 samples were the exact same subvariant as the primary case, implying an intra-household correlation of subvariants of 100%.

#### 9.3 Robustness of main results

This section provides additional analyses to investigate the robustness of our main results. Model I in Tables 12 and 13 provides the odds ratio (OR) estimates of our main model specification. In model II, we only include households with primary cases identified in the period 5-11 January 2022, in order to exclude the atypical transmission patterns between Christmas 2021 and New Year’s Eve 2021/22. In model III, we only include households, where the primary cases have been identified by TestCenter Denmark to account for the potential sampling bias from the healthcare track (Section 6.2). In model IV, we only include households with two persons to account for the natural weighting bias from different sizes of households. In model V, we exclude households, where the primary case was below 10 years of age. In model VI, we exclude households that have previously been infected (defined by having a positive RT-PCR test), to exclude possible hybrid immunity (i.e. immunity from both vaccination and previous infection). In model VII, we only include positive secondary cases that tested positive on day 2-7 (in model I, this window is 1-7), as these could potentially be misclassified co-primary cases. Similarly, in model VIII, we only include positive secondary cases that tested positive on day 3-7. In model IX, we additionally adjust for the primary case sample Ct value, as differences in the viral load could potentially affect the results.

**Table 12:**
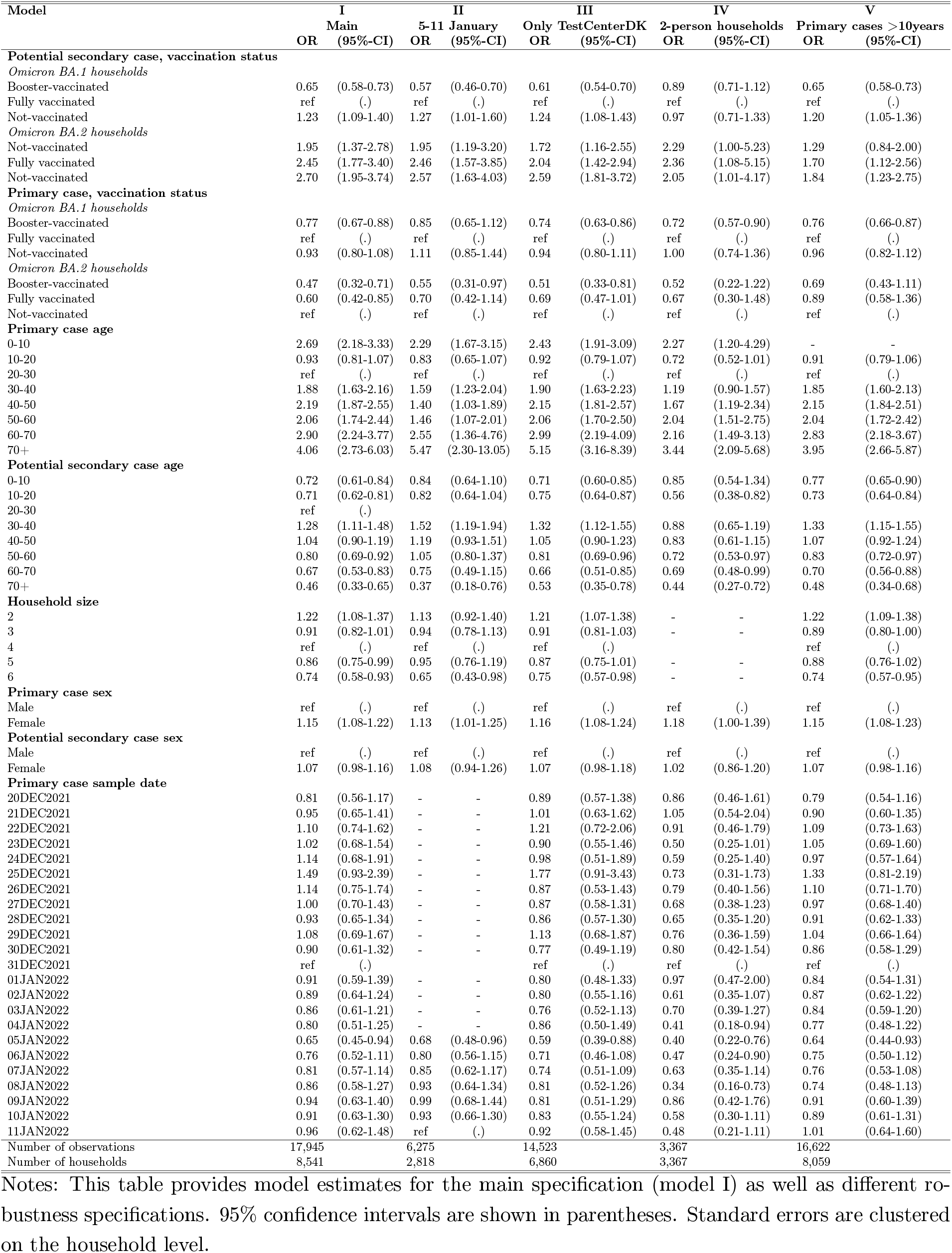
Robustness I

**Table 13:**
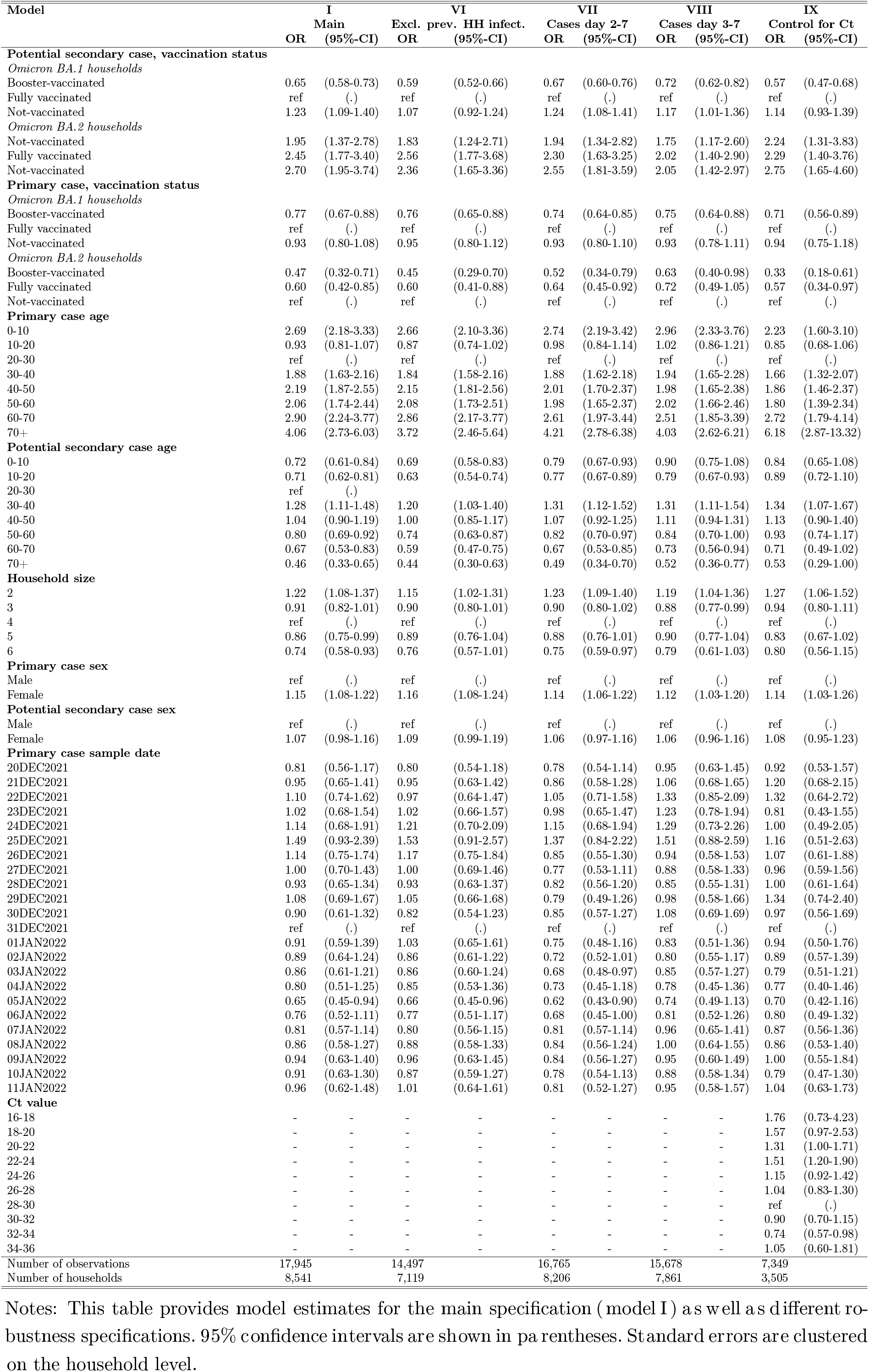
Robustness II

To investigate the sensitivity of our results presented in Table 3, we estimated our model by stratifying the sample (Table 14).

**Table 14:** Relative effect of Omicron BA.2 vs. BA.1, by stratification

|  | Susceptibility |  |  | Transmissibility |  |  |
| --- | --- | --- | --- | --- | --- | --- |
|  | Unvaccinated | Fully vaccinated | Booster vaccinated | Unvaccinated | Fully vaccinated | Booster vaccinated |
| Omicron BA.2 households | 2.74<br>(1.76-4.27) | 1.99<br>(1.26-3.16) | 3.54<br>(1.86-6.76) | 3.92<br>(2.49-6.16) | 1.59<br>(1.10-2.29) | 1.11<br>(0.58-2.14) |
| Omicron BA.1 households | ref<br>(.) | ref<br>(.) | ref<br>(.) | ref<br>(.) | ref<br>(.) | ref<br>(.) |
| Number of observations | 4,631 | 7,838 | 5,476 | 3,500 | 10,659 | 3,786 |
| Number of households | 3,030 | 5,391 | 4,084 | 1,465 | 4,895 | 2,181 |

Notes: This table provides model estimates similar to Table 3, but with stratification of the sample. For the susceptibility estimates, we stratify by the vaccination status of the potential secondary case. For the transmissibility estimates, we stratify by the vaccination status of the primary case. 95% confidence intervals are shown in parentheses. Standard errors are clustered on the household level.

## References

Bager, P., Wohlfahrt, J., Bhatt, S., Edslev, S. M., Sieber, R. N., Ingham, A. C., Stegger, M., Legarth, R., Holten Møller, C., Skov, R. L., Valentiner-Branth, P., Overvad, M., Gram, M. A., Lomholt, F. K., Hallundbæk, L., Espensen, C. H., Gubbels, S. M., Voldst-edlund, M., Karakis, M., Møller, K. L., Olsen, S. S., Fischer, T. K., Barrella Harboe, Z., Johannesen, C. K., van Wijhe, M., Holler, J. G., Simonsen, L., Dessau, R. B. C., Friis, M. B., Fuglsang-Damgaard, D., Pinholt, M., Kirkby, N. S., Thomsen, M. K., Syden-ham, T. V., Coia, J. E., Marmolin, E. S., Fomsgaard, A., Fonager, J., Rasmussen, M., Spiess, K., Marving, E., Cohen, A., Larsen, N. B., Lillebaek, T., Ullum, H., Mølbak, K., & Grove Krause, T. (2022). Reduced Risk of Hospitalisation Associated With Infection With SARS-CoV-2 Omicron Relative to Delta: A Danish Cohort Study. SSRN.

Bomze, D., Sprecher, E., & Gamzu, R. (2021). Effect of a nationwide booster vaccine rollout in Israel on SARS-CoV-2 infection and severe illness in young adults. Travel medicine and infectious disease, 44, 102195.

Chowdhury, S., Bappy, M. H., Chowdhury, S., Chowdhury, M. S., & Chowdhury, N. S. (2022). Omicron Variant (B. 1.1. 529) of SARS-CoV-2, A Worldwide Public Health Emergency! European Journal of Clinical Medicine, 3(1), 5–9.

Darwin, C. (1859). On the Origin of Species by Means of Natural Selection. London: John Murray.

Ferguson, N., Ghani, A., Cori, A., Hogan, A., Hinsley, W., & Volz, E. (2021). Report 49: Growth, population distribution and immune escape of Omicron in England. Imperial College London (16-12-2021), doi: https://doiorg/1025561, 93038.

Jakobsen, K. K., Schmidt Jensen, J., Todsen, T., Kirkby, N., Lippert, F., Vangsted, A.-M., Klokker, M., & von Buchwald, C. (2021). Accuracy of anterior nasal swab rapid antigen tests compared with RT-PCR for massive SARS-CoV-2 screening in low prevalence population. APMIS.

Krause, T. G., Jakobsen, S., Haarh, M., & Mølbak, K. (2012). The Danish vaccination register. Eurosurveillance, 17(17), 20155.

Levine-Tiefenbrun, M., Yelin, I., Katz, R., Herzel, E., Golan, Z., Schreiber, L., Wolf, T., Nadler, V., Ben-Tov, A., Kuint, J., et al. (2021). Decreased SARS-CoV-2 viral load following vaccination. MedRxiv.

Li, H. (2011). A statistical framework for SNP calling, mutation discovery, association mapping and population genetical parameter estimation from sequencing data. Bioin-formatics, 27(21), 2987–2993.

Lyngse, F. P., Mølbak, K., Denwood, M., Christiansen, L. E., Møller, C. H., Rasmussen, M., Cohen, A. S., Stegger, M., Fonager, J., Sieber, R., et al. (2022). Effect of Vaccination on Household Transmission of SARS-CoV-2 Delta VOC. medRxiv.

Lyngse, F. P., Mølbak, K., Skov, R. L., Christiansen, L. E., Mortensen, L. H., Albertsen, M., Møller, C. H., Krause, T. G., Rasmussen, M., Michaelsen, T. Y., et al. (2021a). Increased transmissibility of SARS-CoV-2 lineage B. 1.1. 7 by age and viral load. Nature Communications, 12(1), 1–8.

Lyngse, F. P., Mortensen, L. H., Denwood, M. J., Christiansen, L. E., Møller, C. H., Skov, R. L., Spiess, K., Fomsgaard, A., Lassaunière, R., Rasmussen, M., Stegger, M., Nielsen, C., Sieber, R. N., Cohen, A. S., Møller, F. T., Overvad, M., Mølbak, K., Krause, T. G., & Kirkeby, C. T. (2021b). SARS-CoV-2 Omicron VOC Transmission in Danish Households. MedRxiv.

Majumdar, S. & Sarkar, R. (2021). Mutational and phylogenetic analyses of the two lineages of the Omicron variant. Journal of medical virology.

Mullen, J. L., Tsueng, G., Latif, A. A., Alkuzweny, M., Cano, M., Haag, E., Zhou Jerry Zeller, M., Hufbauer, E., Matteson, N., Andersen, K. G., Wu, C., Su, A. I., Gangavarapu, K., Hughes, L. D., & for Viral Systems Biology outbreak, C. (2022). SARS-CoV-2 (hCoV-19) Mutation Reports.

O’Toole, Á., Hill, V., Pybus, O. G., Watts, A., Bogoch, I. I., Khan, K., Messina, J. P., Covid, T., et al. (2021). Tracking the international spread of SARS-CoV-2 lineages B. 1.1. 7 and B. 1.351/501Y-V2. Wellcome Open Research, 6.

Pearson, C. A., Silal, S. P., Li, M. W., Dushoff, J., Bolker, B. M., Abbott, S., van Schalkwyk, C., Davies, N. G., Barnard, R. C., Edmunds, W. J., et al. (2021). Bounding the levels of transmissibility & immune evasion of the Omicron variant in South Africa. medRxiv.

Pfizer (2021). Pfizer and Biontech submit request to amend U.S. FDA emergency use authorization of their Covid-19 vaccine booster to include all individuals 18 and older) https://cdn.pfizer.com/pfizercom/2021-11/Booster_10K_Efficacy_EUA_Submission_Statement_Final_11921.pdf?linkId=139453336 (accessed 2021-12-21).

Planas, D., Saunders, N., Maes, P., Guivel-Benhassine, F., Planchais, C., Buchrieser, J., Bolland, W.-H., Porrot, F., Staropoli, I., Lemoine, F., et al. (2021). Considerable escape of SARS-CoV-2 Omicron to antibody neutralization. Nature, (pp. 1–7).

Public Health England (2022). SARS-CoV-2 variants of concern and variants under in-vestigation in England. technical briefing 34.

Puhach, O., Adea, K., Hulo, N., Sattonnet-Roche, P., Genecand, C., Iten, A., Bausch, F. J., Kaiser, L., Vetter, P., Eckerle, I., et al. (2022). Infectious viral load in unvaccinated and vaccinated patients infected with SARS-CoV-2 WT, Delta and Omicron. medRxiv.

R Core Team (2021). R: A Language and Environment for Statistical Computing. R Foundation for Statistical Computing, Vienna, Austria.

Ritchie, H. & Roser, M. (2020). Share of SARS-CoV-2 sequences that are the omicron variant, Jan 21, 2022. https://ourworldindata.org/grapher/covid-cases-omicron?country=GBRFRABELDEUITAESPUSAZAFBWAAUS.

Schønning, K., Dessau, R. B., Jensen, T. G., Thorsen, N. M., Wiuff, C., Nielsen, L., Gubbels, S., Denwood, M., Thygesen, U. H., Christensen, L. E., Møller, C. H., Møller, J. K., Ellermann-Eriksen, S., Østergaard, C., Lam, J. U. H., Abushalleeh, N., Meaidi, M., Olsen, S., Mølbak, K., & Voldstedlund, M. (2021). Electronic reporting of diag-nostic laboratory test results from all healthcare sectors is a cornerstone of national preparedness and control of COVID-19 in Denmark. APMIS, 129(7), 438–451.

Spiess, K., Gunalan, V., Marving, E., Nielsen, S. H., Joergensen, M. G., Fomsgaard, A. S., Nielsen, L., Alfaro-Nunez, A., Karst, S. M., Mortensen, S., et al. (2021). Rapid surveillance platforms for key SARS-CoV-2 mutations in Denmark. medRxiv.

SSI (2021). Covid-19 Vaccinedashboard, (accessed 2021-12-22). https://experience.arcgis.com/experience/9824b03b114244348ef0b10f69f490b4/page/Regionalt/.

SSI (2022). Now, an Omicron variant, BA.2, accounts for almost half of all Danish Omicron-cases.

Sundhedsministeriet (2022). Alle restriktioner udløber d. 31. januar.

Wolter, N., Jassat, W., Walaza, S., Welch, R., Moultrie, H., Groome, M., Amoako, D. G., Everatt, J., Bhiman, J. N., Scheepers, C., et al. (2022). Early assessment of the clinical severity of the SARS-CoV-2 omicron variant in South Africa: a data linkage study. The Lancet.

Zhang, X., Wu, S., Wu, B., Yang, Q., Chen, A., Li, Y., Zhang, Y., Pan, T., Zhang, H., & He, X. (2021). SARS-CoV-2 Omicron strain exhibits potent capabilities for immune evasion and viral entrance. Signal transduction and targeted therapy, 6(1), 1–3.

